# Estimation of incidence from aggregated current status data without differential mortality

**DOI:** 10.1101/2021.05.28.21258004

**Authors:** Ralph Brinks

**Affiliations:** Chair for Medical Biometry and Epidemiology, Witten/Herdecke University, Faculty of Health/School of Medicine, D-58448 Witten, Germany; Institute for Biometry and Epidemiology, German Diabetes Center, D-40225 Düsseldorf, Germany; Department for Statistics, Ludwig-Maximilians-University Munich, D-80539 München, Germany

**Keywords:** prevalence, chronic diseases, illness-death model, MCMC estimation, maximum likelihood estimation

## Abstract

We use a historical data about breathlessness in British coal miners to compare two methods centered around a differential equation for deriving the age-specific incidence from aggregated current status data with age information, i.e. age-specific prevalence data. Special focus is put on estimating confidence bounds. For this, we derive a maximum likelihood (ML) estimator for estimating the age-specific incidence from the prevalence data and confidence bounds are calculated based on classical ML theory. Second, we construct a Markov-Chain-Monte-Carlo (MCMC) algorithm to estimate confidence bounds, which implements a weighted version of the differential equation into the prior of the MCMC algorithm. The confidence bounds for both methods are compared and it turns out that the MCMC estimates approach the ML estimates if the prior gives strong weight to the differential equation.

## Introduction

The question about relations between incidence and prevalence of diseases dates back at least to 1934, when Muench examined if the age-specific incidence of yellow fever in Southern America can be reconstructed from a cross-sectional sample about the age-specific prevalence [Mue34]. Muench used the phrase *catalytic curve*, which still found in recent textbooks about infectious disease epidemiology [Vyn10]. In the statistical literature, the term *current status data* is more frequently used than catalytic models. By current status data, it is assumed to have a cross-sectional sample where for each study participant, current age and current disease status are known. A literature review about current status data is found in [McK11].

In epidemiology, current status data are often aggregated in different age groups. For example, the data in Muench’s article about yellow fever in Colombia is given as positive cases from a number of tests applied in five age groups: 5-9, 10-14, 15-19, 20-39 and 40+ (Table 1 in [Mue34]).

**Table 1:** Data about breathlessness in British coal miners [Ela14].

| Age group<br>$k$ (in years) | Number<br>observed ($n_k$) | Number with<br>condition ($c_k$) | Prevalence<br>($p_k$) (%) |
| --- | --- | --- | --- |
| 20-24 | 1952 | 16 | 0.820 |
| 25-29 | 1791 | 32 | 1.787 |
| 30-34 | 2113 | 73 | 3.455 |
| 35-39 | 2783 | 169 | 6.073 |
| 40-44 | 2274 | 223 | 9.807 |
| 45-49 | 2393 | 357 | 14.92 |
| 50-54 | 2090 | 521 | 24.93 |
| 55-59 | 1750 | 558 | 31.89 |
| 60-64 | 1136 | 478 | 42.08 |

In this technical note, we use a historical data about breathlessness in British coal miners [Ela14] to compare three methods centered around a differential equation for deriving the age-specific incidence from aggregated current status data. Using the differential equation for estimating incidence from prevalence data has been described in [Bri13] and has been compared to other methods in [Lan16]. In this work, special focus is put on estimating confidence bounds based on the differential equation. The data we use here are shown in Table 1.

First, we derive a novel maximum likelihood (ML) estimator for inferring the age-specific incidence from the prevalence data in Table 1. Confidence bounds are calculated from classical ML theory. In a second approach, we construct an Markov-Chain-Monte Carlo (MCMC) algorithm to estimate confidence bounds for the incidence estimates. Finally, the confidence bounds for both methods are compared.

### Estimation of the age-specific incidence

Recently, we have shown that the age-specific prevalence *p* of a chronic condition at some time *t* is related to the age-specific incidence density *i* (synonym: incidence rate), and mortality rates *m*_0_, and *m*_1_ via a partial differential equation (PDE) [Bri14]. Figure 1 shows the underlying multi-state model with the possible transitions and associated rates.

**Figure 1:**
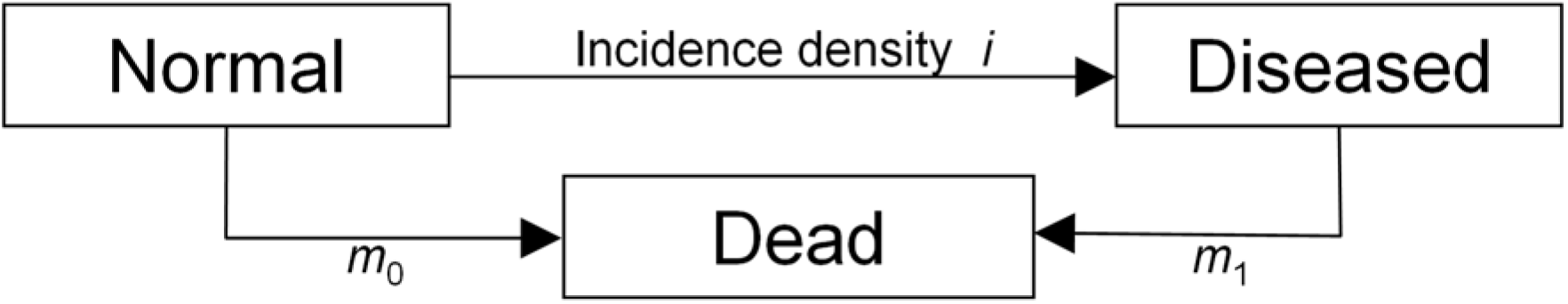
Illness-death model for a chronic condition (‘Diseased’) and associated transition densities: incidence density *i*, mortality without (*m*_0_) and with the disease (*m*_1_).

We consider the case without migration and without differential mortality (i.e., *m*_0_ = *m*_1_), where the PDE from [Bri14] reads as

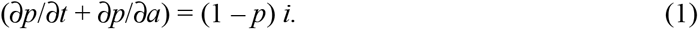

If we assume furthermore, that the incidence density *i* in Equation (1) does not depend on time *t*, the PDE (1) becomes an ordinary differential equation (ODE)

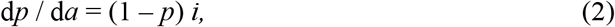

which has the general solution

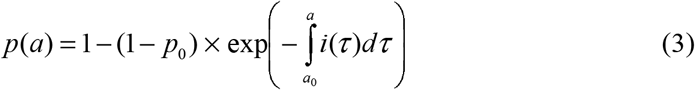

where *p*_0_ = *p*(*a*_0_) is the initial condition.

Equation (2) is the basis for a straightforward estimator of the age-specific incidence density *i*. If we can estimate the derivative d*p* from the age-specific prevalence *p*, we obtain *i* = d*p*/(1 – *p*).

Example 1: We fit a linear model logit(*p*(*a*)) = log(*p*(*a*)/{1 – *p*(*a*)}) = β_0_ + β_1_ ⨯ *a* at the midpoints of the age groups *a* = 22.5, 27.5, …, 62.5 to the data from Table 1. We obtain β_0_ = –7.02 and β_1_ = 0.110. Inserting *p*(*a*) = expit(β_0_ + β_1_ × *a*) into *i* = d*p*/(1 – *p*) yields *i*(*a*) = β_1_ *p*(*a*). The expit-function is the inverse of the logit-function, i.e., expit = exp/(1+exp), and has the derivative expit′ = expit × (1 – expit). The associated age-specific incidence *i*(*a*) = β_1_ expit(β_0_ + β_1_ × *a*) with β_0_ = –7.02 and β_1_ = 0.110 is plotted as black line in Figure 2. For comparison, the blue line shows the estimates [Ela14] has given. Note the negative estimate at age 57.5 from [Ela14].

**Figure 2:**
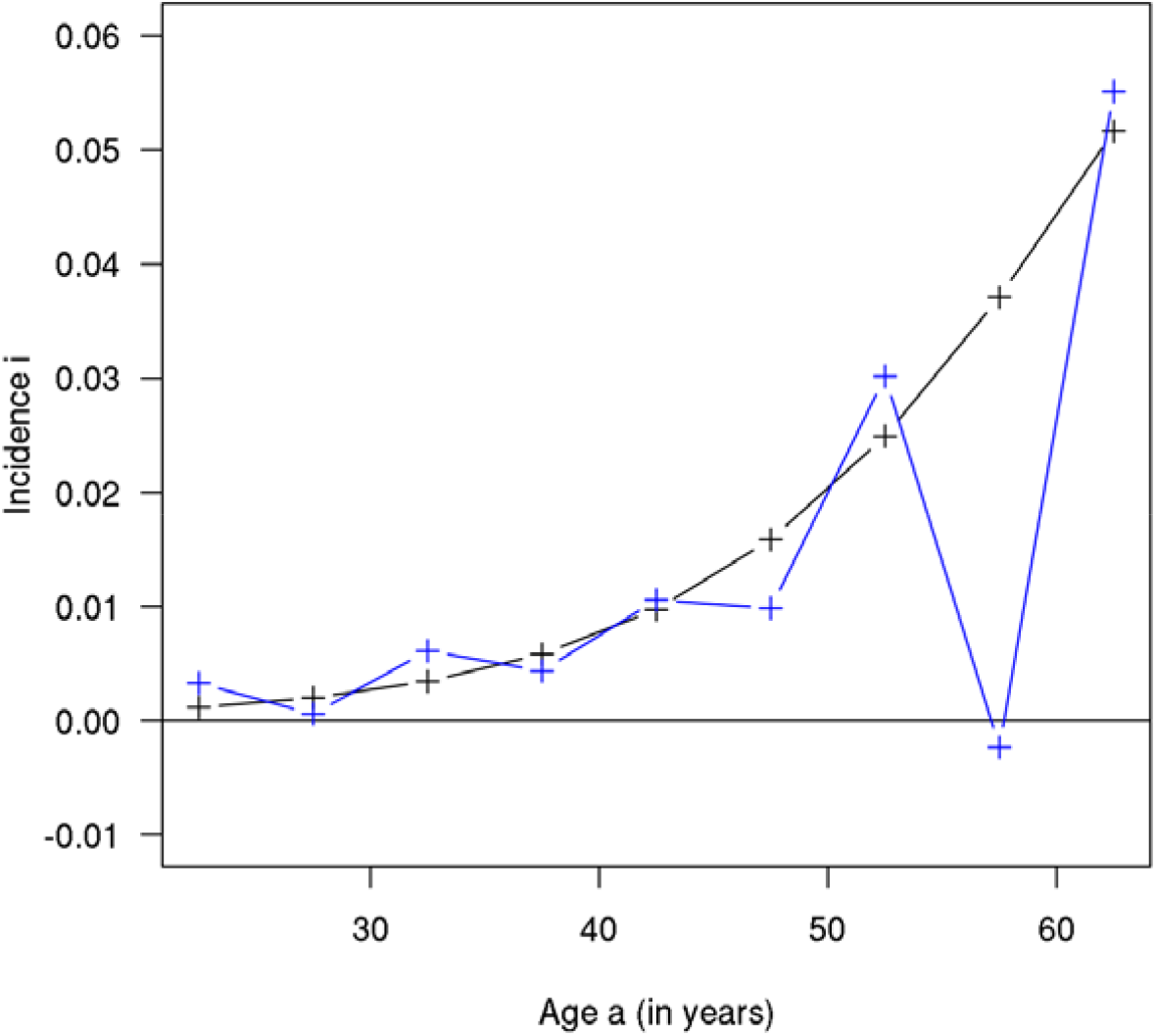
Estimated age-specific incidence for the data in Table 1 as calculated in Example 1.

Example 1 has demonstrated how the theory of differential equations can be used to estimate the age-specific incidence from aggregated current status data (prevalence data) of a chronic condition. In this work, we are interested in estimating confidence bounds for the age-specific incidence.

### Maximum likelihood estimation

The binomial likelihood function *L* for the aggregated current status data, like the data in Table 1, is given by

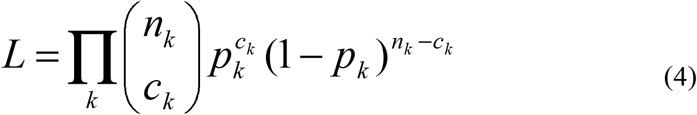

where *k* is the index for the age group, *n*_k_ and *c*_k_ are the numbers of subjects and cases in an age group indexed by *k*, respectively. *p*_k_ = *c*_k_/*n*_k_ is the fraction of subjects in age group *k* with the condition under consideration (prevalence). In Table 1 we have nine age groups. Hence, for these data the product in Equation (4) consists of nine factors.

With a view to Figure 2, we have hints that the age-specific incidence density *i*(*a*) grows exponentially with age. Hence, we make the approach *i*(*a*) = exp(γ_0_ + γ_1_ × *a*) with coefficients γ_0_, γ_1_. If we substitute this incidence into Equation (3) with the initial condition *a*_0_ = 20 and *p*(20) = *p*_0_ = 0, we obtain *p*(*a*) = 1 – exp(*h*(20) – (*a*)) with *h*(*z*) = exp(γ_0_ + γ_1_ × *z*)/γ_1_. Then, this *p* is evaluated at the age group midpoints *a*_k_ = 22.5, …, 62.5 and finally substituted into Equation (4). We end up at a likelihood function *L* = *L*(γ_0_, γ_1_), which by optimization can be used to calculate a maximum likelihood estimator for both coefficients, γ_0_ and γ_1_. If we additionally calculate the 95% confidence intervals using the inverse of the Fisher information matrix for large sample approximation of the variance-covariance matrix [Woo15], we obtain the results shown in Table 2.

**Table 2:** Maximum likelihood estimators for the coefficients γ_0_ and γ_1_used for parameterization of the age-specific incidence.

|  | Point estimate | 95% confidence bounds |
| --- | --- | --- |
| $\gamma_0$ | -7.8237 | -8.0590 to -7.5883 |
| $\gamma_1$ | 0.07559 | 0.07006 to 0.08111 |

### MCMC approach

For data resulting from a PDE data generating process *F*, Xun and colleagues had the idea to separate the model fit *g* = *g*(θ) with unknown parameters θ to the observed data *Y* on the one hand and the model’s accordance to the PDE *F* on the other [Xun13]. Hence, we have a functional

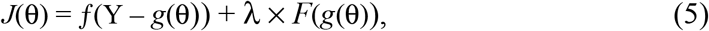

with a relaxation parameter λ > 0, which penalizes deviations from the data generating PDE model *F*. Xun et al. have chosen the function *f* for the data fit to be the square function *f* (*Y* – *g*(θ)) = {*Y* – *g*(θ)}^2^. Other choices are possible, and we will use a weighted square approach below.

In our research question, the observed data *Y* is the age-specific prevalence *p* in Table 1. As in Example 1, we fit the *p* by an expit model: *p*(*a*) = expit(β_0_ + β_1_ × *a*). In addition, we fit the incidence *i* to an exponential model: *i*(*a*) = exp(γ_0_ + γ_1_ × *a*). The function *F* describing the ODE (3) is defined via a vector of parameters to be estimated θ = (β_0_, β_1_, γ_0_, γ_1_):

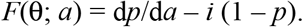

Here, we have *F*(θ; *a*) = {β_1_ expit(β_0_ + β_1_ × *a*) – exp(γ_0_ + γ_1_ × *a*)} {1 – expit(β_0_ + β_1_ × *a*)}. *F* is squared and integrated over the whole age range, in case of the data in Table 1 from *a*_0_ = 20 to *a*_e_ = 65 (years):

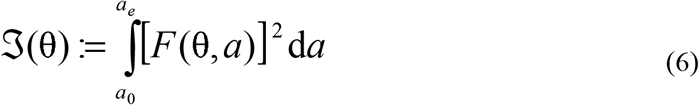

If we have a good agreement between the estimated parameters θ and the data generating differential equation (2), ℑ(θ) as defined in Equation (5) is close to zero. We set up a MCMC algorithm to estimate θ and use ℑ(θ) as a prior [Woo15]. To control the relative importance, we multiply with an penalization factor λ > 0. This leads us to following prior ∝ exp{– λ ℑ(θ)}. For the log-likelihood in the MCMC algorithm, we choose

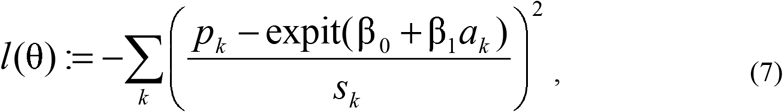

with *s*_k_ = *p*_k_ (1 – *p*_k_)/*n*_k_ which corresponds to a weighted least squares approach. Again, the *a*_k_ in Equation (7) are the midpoints of the age-groups in Table 1.

Given the prior and the log-likelihood, we are able to run a MCMC scheme based on the Metropolis-Hastings method [Woo15]. For each choice of λ in the range 1000 to 2,000,000, we use 500,000 iterations and discard the first 200,000 iterations as burn-in period.

The role of the relaxation parameter λ is shown in Figure 3, where the parameters γ_0_ (left panel) and γ_1_ (right panel) with 95% confidence bounds as estimated by the MCMC algorithm are plotted over the value λ. For comparison with the ML estimates and their confidence bounds from Table 2, the dashed blue lines and the blue areas represent the point estimates and confidence bounds for γ_0_ and γ_1_, respectively.

**Figure 3:**
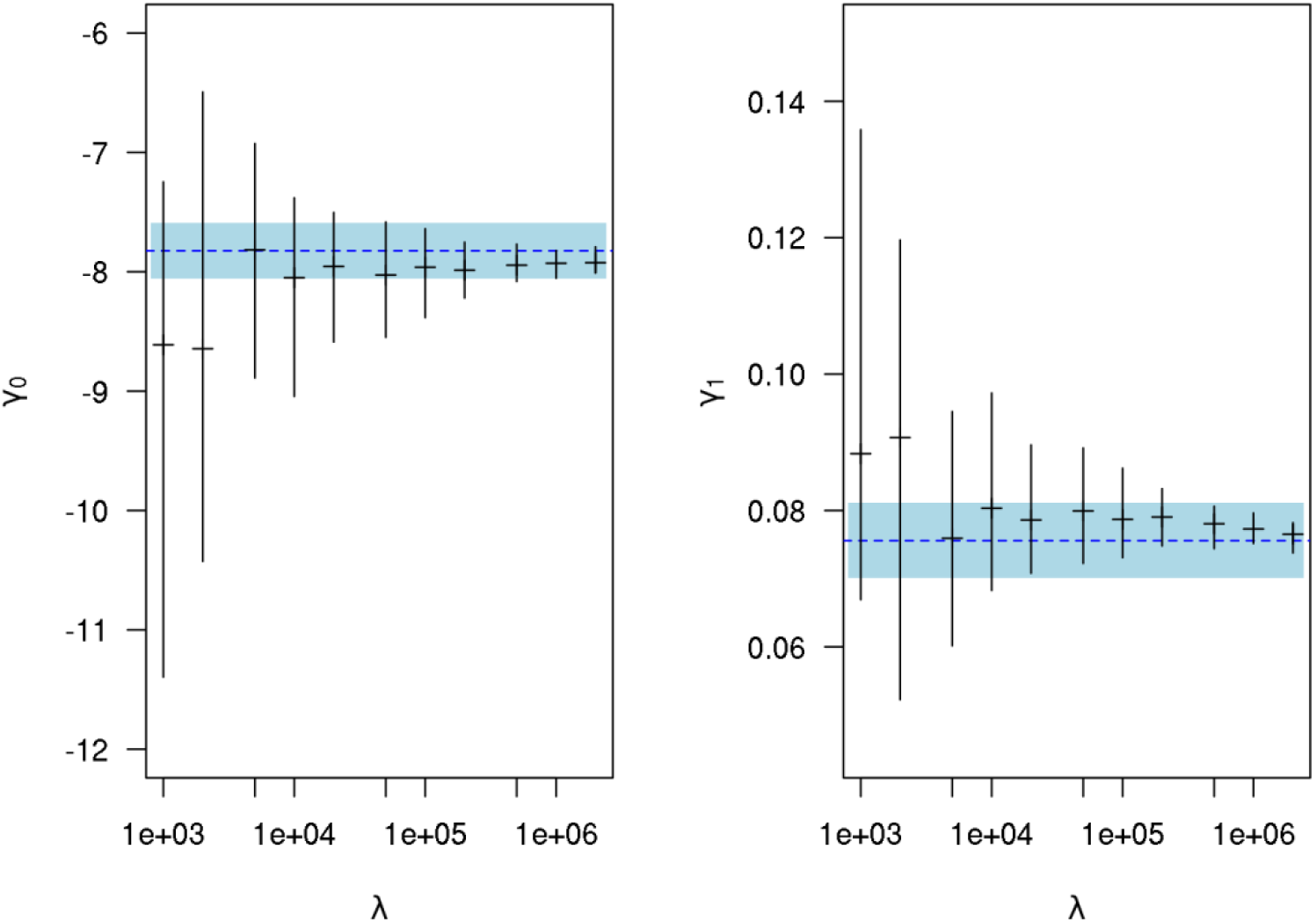
Incidence parameters γ_0_ (left panel) and γ_1_ (right) with 95% confidence bounds (vertical bars) as calculated by the MCMC algorithm with different weighting parameters λ (abscissa). The blue dashed lines and blue areas represent the ML estimates together with the 95% confidence bounds from Table 2.

Interestingly, as the parameter λ increases, the length of the confidence bounds from the MCMC decreases and finally lie inside the confidence bounds estimated from the ML estimator.

## Discussion

In this work, we have described two methods about statistical inference around a differential equation, which relates the age-specific prevalence of a chronic condition with the underlying age-specific incidence.

The differential equation is not new, early approaches heading into a similar direction go back at least to the 1990ies [Bru99]. The innovative aspects in this work are twofold: 1) the link with standard maximum likelihood (ML) inference and 2) the implementation of the differential equation into the prior of an MCMC algorithm. While the maximum likelihood approach seems rather straightforward, the idea about MCMC goes back to rather recent work of Xun and colleagues [Xun13]. We could demonstrate that both methods are related, for example by the observation that the MCMC confidence bounds tend to the ML confidence bounds if the relaxation parameter λ is chosen sufficiently large. So far this is only an observation from a computational experiment, which of course requires a deeper theoretical investigation about the relation of ML theory and MCMC simulations.

These two methods, ML and MCMC estimation, in combination with differential equations like Eq. (1) extend the methodological toolbox in epidemiology as requested about a decade ago [Chu10]. Until now, we had to employ re-sampling techniques to obtain confidence bounds in the context of differential equations. In re-sampling, a number of random samples from the reported distributions of the input parameters have been drawn to estimate how the uncertainty in the input parameters propagate through the differential equations into the outcomes. An example for the this type of re-sampling is described, for example, in [Bri15].

## Data Availability

The source codes including the data sets for the analysis (and preparation of graphs) underlying this paper and for running the MCMC algorithm are available in the open public repository Zenodo. The links and DOIs are given in the manuscript.

https://doi.org/10.5281/zenodo.4816749

https://doi.org/10.5281/zenodo.4770055

## Appendices

## Data availability

The source codes including the data sets for the analysis (and preparation of graphs) underlying this paper [Bri21a] and for running the MCMC algorithm [Bri21b] are available in the open public repository Zenodo.

## Acknowledgements

The author gratefully acknowledges the Gauss Centre for Supercomputing e.V. (www.gauss-centre.eu) for funding this project by providing computing time on the GCS Supercomputer SuperMUC at the Leibniz Supercomputing Centre (www.lrz.de).

## References

[Bri13] Brinks R, Landwehr S, Icks A, Koch M, Giani G. Deriving age-specific incidence from prevalence with an ordinary differential equation. Stat Med. 30;32(12):2070–8, 2013 doi: 10.1002/sim.5651.

[Bri14] Brinks R, Landwehr S: Age-and time-dependent model of the prevalence of non-communicable diseases and application to dementia in Germany. Theoretical Population Biology, 92:62–68, 2014

[Bri15] Brinks R, Hoyer A, Kuss O, Rathmann W: Projected Effect of Increased Active Travel in German Urban Regions on the Risk of Type 2 Diabetes. PLOS ONE 10(4): e0122145, 2015. https://doi.org/10.1371/journal.pone.0122145

[Bri21a] Brinks R: Statistical Analysis for Skinfaxi Project, Zenodo repository DOI 10.5281/zenodo.4816749 https://doi.org/10.5281/zenodo.4816749

[Bri21b] Brinks R: Estimation of age-specific incidence from prevalence: novel MCMC approach, Zenodo repository DOI 10.5281/zenodo.4770055 https://doi.org/10.5281/zenodo.4770055

[Bru99] Brunet RC, Struchiner CJ: A non-parametric method for the reconstruction of age- and time-dependent incidence from the prevalence data of irreversible diseases with differential mortality. Theoretical Population Biology 56(1): 76–90, 1999.

[Chu10] Chubb MC, Jacobsen KH: Mathematical modeling and the epidemiological research process. European Journal of Epidemiology, 25(1), 13–19, 2010

[Ela14] Elandt-Johnson RC, Johnson NL: Survival models and data analysis. John Wiley & Sons, 2014

[Lan16] Landwehr S, Brinks R. A comparative study of prevalence-based incidence estimation techniques with application to dementia data in Germany. Stat Med 28;35(5):768–81, 2016, DOI: 10.1002/sim.6736

[McK11] McKeown KM: Topics in Current Status Data, PhD Dissertation at the University of Berkley, 2011

[Mue34] Muench H: Derivation of Rates from Summation Data by the Catalytic Curve, Journal of the American Statistical Association 29 (185):25–38, 1934

[Vyn10] Vynnycky E, White RG: An Introduction to Infectious Disease Modelling, Oxford University Press, 2010

[Woo15] Wood SN, Core Statistics, Cambridge University Press, 2015

[Xun13] Xun X, Cao J, Mallick B, Maity A, Carroll RJ: Parameter estimation of partial differential equation models. Journal of the American Statistical Association, 108(503), 1009–1020, 2013

